# Subjective mental health and need for care among psychiatric outpatients during the COVID-19 pandemic: results from an outreach initiative in Sweden

**DOI:** 10.1101/2020.11.10.20229039

**Authors:** Oskar Flygare, Volen Z. Ivanov, Roland Säll, Henrik Malaise, Christian Rück, Nitya Jayaram-Lindström, Lina Martinsson

## Abstract

**Importance:** The ongoing COVID-19 pandemic restricts access to care for psychiatric patients. The physical and mental well-being of patients with severe mental illness in the current circumstances is unknown.

**Objective:** To evaluate physical and mental well-being, subjective mental health, and need for updated psychiatric management plans in a sample of patients with severe mental illness during the early stages of the COVID-19 pandemic.

**Design:** Cross-sectional study of structured telephone assessments conducted between April 23 and June 30, 2020.

**Setting:** Regional psychiatric outpatient care centre in Stockholm, Sweden.

**Participants:** Patients who had not been in contact with their psychiatric clinic between April 9 and April 23, 2020. A total of 1071 patients were contacted by phone.

**Exposures:** Occurrence of respiratory symptoms, changes in psychiatric symptoms, and the need for updated psychiatric management plans, as determined by the telephone assessors. Subjective mental health rated 0-100 by patients.

**Main Outcomes and Measures:** Self-rated physical, respiratory and psychiatric symptoms according to a semi-structured interview. Subjective mental health rated on a scale from 0-100.

**Results:** Patients (n = 1071) were on average 45 years old (SD = 16.9), of which 570 (53%) were female. Neurodevelopmental disorders, psychotic disorders, and bipolar disorder were the most common diagnostic categories. The majority of respondents reported no respiratory symptoms (86%), and few reported light (10%) or severe (4%) respiratory symptoms. Similarly, most patients reported no worsening in psychiatric symptoms (81%). For those who reported a worsening of psychiatric symptoms (19%), the psychiatric management plans that were already in place were deemed appropriate in most cases (16.5%), whereas 22 patients (2.5%) reported a worsening of psychiatric symptoms that warranted an earlier or immediate follow-up by their psychiatric clinic. Patients rated their subjective mental health on a 0-100 scale as 70.5 [95% CI 69 - 71.9] on average (n = 841). Response rates to the questions of the structured assessment varied from 79% - 82%.

**Conclusions and Relevance:** The majority of patients reported no respiratory symptoms, no change in psychiatric symptoms and a rather high subjective well-being. Patients in psychiatric care with a mental health care plan experienced stability in the management of their psychiatric symptoms and general well-being, and only a minority were in need of acute support during the early pandemic phase in Stockholm, Sweden.

**Key Points:** *Question:* What is the physical and mental health of patients with severe mental illness during the early phase of the COVID-19 pandemic?

*Findings:* In this cross-sectional study that included 1071 patients at a psychiatric outpatient clinic, the proportion of patients reporting respiratory symptoms were 4%. In addition, 19% of patients reported a worsening of psychiatric symptoms, with 2.5% needing an earlier follow-up than was planned.

*Meaning:* Patients with severe mental illness experienced stability in the management of their psychiatric symptoms during the early pandemic phase in Sweden.

## Background

The ongoing COVID-19 pandemic has wide-ranging consequences for everyday life across the globe, with physical distancing, closing of public spaces and other measures^1^ taken by most countries to limit the spread of the virus^2^. These restrictions also affect patients with severe mental illness (SMI) due to the rapid and dramatic changes that took place in the landscape of regular psychiatric health care. Public health experts and researchers have warned that the impact of the pandemic in itself, societal measures to limit the spread of disease, and changes in health care delivery may create a perfect storm of secondary adverse consequences, such as increase in suicide risk, however such an increase has not been observed in available data to date^3,4^. Patients with SMI are a particularly vulnerable group in society^5^, and recent findings also indicate that they are more likely to contract and die from COVID-19 compared to the general population^6,7^. Policy makers and funders now call for studies^8^ that can shed light on the effects of the COVID-19 pandemic on vulnerable groups, and to improve knowledge on how to mitigate the negative effects of the pandemic.

Patients with SMI may experience a worsening of symptoms as a consequence of strict social distancing measures, economic downturns and changes in the delivery of care^9^. Reports emerging from China indicate that home isolation has negative psychological effects^10^ for a majority of the population^11^. In response to the disease outbreak, mental health services in the Lombardy region in Italy^12^ restricted access to day centres and greatly reduced the number of home-visits. A Chinese survey on 2065 psychiatric outpatients reported that 21% of respondents experienced a deterioration of their mental health, and 22% reported a lack of access to adequate care during the pandemic^13^. Patients may also be hesitant to contact or visit their psychiatric clinic out of fear of contamination or putting an undue burden on the health care system. At the same time, they are faced with a situation of having to access and navigate care when needed via remote and digital modes^14^, a method that is not only novel but also raises the threshold of difficulty, for many in this already vulnerable group. For example, there was a 25% decrease in visits to the emergency psychiatric unit in Stockholm between March 1st and April 23rd, compared to the same dates in 2019. Taken together, these reports on possible negative impact in patients with SMI warrants research that investigates the consequences of the COVID-19 pandemic on the psychiatric population.

The aims of this project were to study the effects of the COVID-19 pandemic on the physical and mental health of patients with SMI at a large psychiatric clinic in Stockholm, Sweden during the early pandemic phase.

## Method

### Study design

This study describes the results of an outreach initiative that was part of the response to the COVID-19 pandemic at a large psychiatric clinic in Stockholm, Sweden. Data was extracted from the electronic health records of each individual, where the outreach phone calls were documented. The study was approved by the Ethical Review Agency in Sweden (no. 2020-03474). The need for informed consent was waived since the research team only accessed pseudonymized data where it was not possible to identify individual patients.

### Procedure

Between April 23rd and June 30th, patients who had not been in touch with their psychiatric clinic since April 9 were contacted via telephone by clinical staff. The purpose was to provide information about available care, to arrange an appointment if deemed necessary, and assess changes in psychiatric symptoms since the beginning of the pandemic in Sweden (March 1st). The assessments contained a series of semi-structured questions about the patients’ overall health, psychiatric symptoms, and reasons for not contacting their psychiatric clinic. Patients were informed about how they could contact their psychiatric clinic, and psychiatric management plans were updated if necessary (for example risk management plans or changes in pharmacological treatment). The phone calls followed a pre-specified routine and staff entered responses according to a template specifically designed for the outreach contact. For respiratory symptoms, the staff could enter one of four options: 1) No respiratory symptoms, 2) Light respiratory symptoms, no need for follow-up in health care, 3) Severe respiratory symptoms, has already been in touch with health care, 4) Severe respiratory symptoms, has not been in touch with health care. For psychiatric symptoms the responses were categorized as follows: 1) No signs of worsening of psychiatric symptoms, 2) Worsening of psychiatric symptoms but existing psychiatric management plan is adequate, 3) Worsening of psychiatric symptoms and need for earlier follow-up, 4) Worsening of psychiatric symptoms and need for immediate follow-up. Staff attempted to contact patients on two consecutive days, and sent a letter with instructions to contact their clinic if they were not reachable by phone.

### Participants

Eligible individuals were patients listed at the psychiatric clinic who had not been in touch with the clinic in the last 14 days prior to April 23, 2020. Patients listed at the psychiatric clinic were either waiting for treatment, had an ongoing treatment, or other scheduled appointments (such as regular visits) in order to follow up the effects of pharmacological and psychological treatments. The psychiatric clinic (Psykiatri Sydväst; Huddinge Karolinska Hospital) is part of the public health care system in Stockholm Region and responsible for providing psychiatric care to residents living in the south-west region of Stockholm, which has 207 454 residents. The clinic is divided into specialized sub-units, each focused on a specific type of diagnosis or care process: emergency psychiatric care, intake and diagnostics, anxiety disorders and depression, psychotic disorders, bipolar disorder, neurodevelopmental disorders, personality disorders, obsessive-compulsive and related disorders, traumatic disorders, and one unit for deaf patients with psychiatric disorders.

### Statistical analyses

The content of the electronic health records was extracted and classified according to pre-specified categories from the outreach template. Data on age and gender was included, and diagnosis was categorized according to the psychiatric unit where the patient was listed. This study did not involve hypothesis testing and the results presented below are descriptive. For the outcomes of physical symptoms, psychiatric symptoms, and changes to psychiatric management plans, the frequency of each type of response was summarized and compared to the total number of responses to obtain percentages. For the rating of subjective health, the mean value and standard deviation was calculated. The responses were compared between the units using linear or logistic regression, including age and gender as co-variates. The statistical software R (version 4.0.2)^15^ was used for all analyses. Scripts used for statistical analyses are available on the Open Science Framework (http://doi.org/10.17605/OSF.IO/NP7MU).

## Results

### Sample characteristics

In total, 1071 patients were contacted via telephone and responses to the specific questions were answered by a majority (79%-82%). The average age of the patients was 45 years (SD = 16.9, range 19-89), and 570 (53%) were women. The majority of the assessments were conducted at the unit for neurodevelopmental disorders (n = 293, 25%), followed by psychotic disorders (n = 249, 21%) and bipolar disorder (n = 245, 21%). Telephone assessments were completed in April (n = 16, 1%), May (n = 807, 75%) and June (n = 248, 23%).

### Occurrence of respiratory symptoms

A total of 879 patients responded to the question about whether they had experienced any respiratory symptoms, of which a majority reported no symptoms (n = 755, 86%). Fewer reported light symptoms that did not warrant contact with health care (n = 87, 10%) or severe symptoms that indicated a need to visit the health care system for assessment and treatment (n = 37, 4%). Light or severe symptoms were less common among men compared to women (OR = 0.5 [95% CI 0.33 to 0.76], SE = 0.22, *p* = 0.001), and were less common at the unit for bipolar disorder compared to other units (OR = 0.21 [95% CI 0.09 to 0.46], SE = 0.41, *p* < 0.001).

### Changes in psychiatric symptoms and management plans

The majority of patients who responded to the question about their psychiatric symptoms (n = 861) reported no immediate worsening of their psychiatric symptoms (n = 696, 81%), and among those who reported a worsening of symptoms (n = 165, 19%) the psychiatric management plan already in place was deemed adequate in most cases (n = 143, 16.6%). A total of 22 patients (2.5%) needed an earlier or immediate follow-up at their psychiatric clinic than was previously planned. Men were less likely to report a worsening of symptoms (OR = 0.51 [95% CI 0.35 to 0.75], SE = 0.19, *p* = 0.001), and patients at the unit for bipolar disorders were less likely to experience a worsening of symptoms compared to the other units (OR = 0.22 [95% CI 0.10 to 0.44], SE = 0.37, *p* < 0.001). In contrast, patients at the emergency psychiatric unit (OR = 2.19 [95% CI 1.29 to 3.74], SE = 0.27, *p* = 0.004) and the unit for anxiety disorders and depression (OR = 1.85 [95% CI 1.02 to 3.32], SE = 0.3, *p* = 0.04) were more likely to report a worsening of their psychiatric symptoms.

### Subjective rating of mental health

Patients were asked to rate their mental health on a scale from 0 (worst possible health) to 100 (best possible health.) The average rating was 70.5 [95% CI 69 - 71.9] among the 841 respondents to this question. Men reported better mental health compared to women (Estimate = 3.73 [95% CI 0.95 to 6.51], SE = 1.41, *p* = 0.008) and there was a positive association between year of birth and subjective mental health (Estimate = 0.11 [95% CI 0.012 to 0.21], SE = 0.05, *p* = 0.029), as younger patients rated their mental health higher on average. Patients at the unit for bipolar disorder (Estimate = 12.5 [8.07 to 16.96], SE = 2.26, *p* < 0.001) and psychotic disorders (Estimate = 4.84 [95% CI 0.59 to 9.09], SE = 2.24, *p* = 0.026) reported a higher subjective mental health, whereas patients at the emergency psychiatric unit (Estimate = −8.35 [95% CI −13.03 to −3.67], SE = 2.38, *p* = <0.001), unit for traumatic disorders (Estimate = −12.09 [95% CI −20.44 to −3.75], SE = 4.25, *p* = 0.005), and unit for anxiety disorders and depression (Estimate = −6.14 [95% CI −11.35 to −0.94], SE = 2.65, *p* = 0.021) rated their mental health as lower.

## Discussion

This study describes the results of a clinical outreach initiative taken at a large psychiatric clinic in Stockholm during the early phase of the COVID 19 pandemic. The need for an outreach initiative was established after observing a dramatic reduction (25%) in the number of patients seeking emergency psychiatric care in Stockholm in March and April 2020, compared to the same period in 2019. The purpose of the outreach was threefold: 1) evaluate the physical and mental health of patients who had not been in contact with their psychiatric clinic during the early pandemic phase, 2) provide emergency response to those who needed it and, 3) inform that care was available to them.

Although 19% of 861 patients had experienced a worsening of their psychiatric symptoms, the existing psychiatric management plans were sufficient in the majority of cases with only 2.5% needing an earlier or immediate follow-up. Interestingly, few patients (4%) reported respiratory symptoms that indicated a need for follow-up in the health care system. The findings also indicate that men, compared to women, reported fewer respiratory symptoms, were less likely to report a worsening of their psychiatric symptoms, and rated their subjective mental health as better. Furthermore, age showed no association with any of the outcomes in this study. Finally, patients at the unit for bipolar disorder responded more positively to all of the questions, likely due to follow-up within psychiatry also in stable phases. This is in contrast to the emergency unit, where patients are only seen in the acute phase of mental illness.

Similar to an early Chinese survey of 2065 psychiatric patients^13^, we found a comparable rate of deterioration in psychiatric symptoms at 20%. However, the access to care was higher in the Swedish sample and fewer patients reported unmet needs in the current study: 22% of respondents in the Chinese survey could not access their routine psychiatric care due to suspended hospital visits, whereas in the present study only 2.5% of patients needed an updated psychiatric management plan. Further, psychiatric wards in Sweden remained open and patients with mental illness as well as those infected with COVID-19 were able to receive inpatient care in isolated wards. An early report from the Lombardy region in Italy describes a closing of psychiatric wards and reallocation of staff towards managing patients with COVID-19^12^, which was not the case in Sweden. Although methodological differences in sampling and screening may explain differences between studies across countries, we speculate that the policies in Sweden, for example no lockdown, and sustained functionality of the specialised psychiatric care may have resulted in better access to care among individuals with severe mental illness during the early pandemic phase^16^.

This study is not without limitations. First, the study population was restricted to those patients who had not been in contact with their psychiatric clinic during the early stages of the COVID-19 pandemic in Sweden. The reasons for not being in touch likely varied between the patients: some may not have been in touch because they had been feeling better after receiving treatment, or were in a stable phase. However, for other patients the reason for not being in touch with their clinic could have been a direct consequence of the pandemic or the societal measures taken to reduce transmission of the virus. Second, there is no data before the pandemic to serve as a reference point. It is likely that, at any given time, some patients will report a deterioration in psychiatric symptoms, for example. Without a direct comparison it is hard to assess whether the findings indicate that the situation has gotten worse or can be viewed as relatively stable. Third, the impact of COVID-19 and the policies implemented to reduce transmission of the virus vary between countries and regions within countries, making it difficult to generalize the current findings to other contexts.

The long-term effects of the COVID-19 pandemic on patients with SMI is currently unknown but there is reason to believe that patients with SMI will be negatively affected. For example, this group of individuals may be particularly vulnerable to the long-term economic effects leading to unemployment and further isolation from their already frail social networks. Further, SMI in itself is associated with an increased risk of COVID-19^6,7^. There is therefore a clear need for continued monitoring of the health in vulnerable populations, such as patients with SMI, as the COVID-19 pandemic continues.

## Conclusions

In conclusion, this study on respiratory symptoms, changes in psychiatric symptoms, and subjective mental health among patients with SMI who had not been in contact with their clinic for two weeks during the early pandemic phase in Stockholm, found that respiratory symptoms requiring follow-up in the healthcare system were rare (4%) and that 19% of patients reported a deterioration in their psychiatric symptoms. However, existing psychiatric management plans were adequate in the majority of cases and few patients needed an early follow-up (2.5%). Men reported fewer respiratory symptoms, lower rates of deterioration in psychiatric symptoms, and better subjective mental health compared to women, and patients at the unit for bipolar disorder–typically treated in stable phases as well as during manic or depressive episodes–responded more positively to all outcomes compared to patients at other units. These findings relate to the early pandemic phase in Sweden, and continued monitoring of vulnerable groups, such as patients with severe mental illness, is needed as the pandemic continues and long-term effects can be observed.

## Supporting information

STROBE checklist

## Data Availability

Requests for additional information should be made to the first or last author. Due to Swedish and European Union data protection and privacy legislation, we generally do not share patient-level data.

https://doi.org/10.17605/OSF.IO/NP7MU

## Authors’ contributions

*Concept and design:* Flygare, Ivanov, Säll, Malaise, Rück, Martinsson.

*Acquisition, analysis, or interpretation of data:* All authors.

*Drafting of the manuscript:* Flygare, Ivanov, Jayaram-Lindström, Martinsson.

*Critical revision of the manuscript for important intellectual content:* All authors.

*Statistical analysis:* Flygare.

*Administrative, technical, or material support:* All authors.

## Additional contributions

We thank the staff of Psykiatri Sydväst who conducted the outreach interviews with patients. We thank Sead Omérov and Mattias Agestam at Region Stockholm for assistance with data preparation and extraction.

---

**Figure 1.**
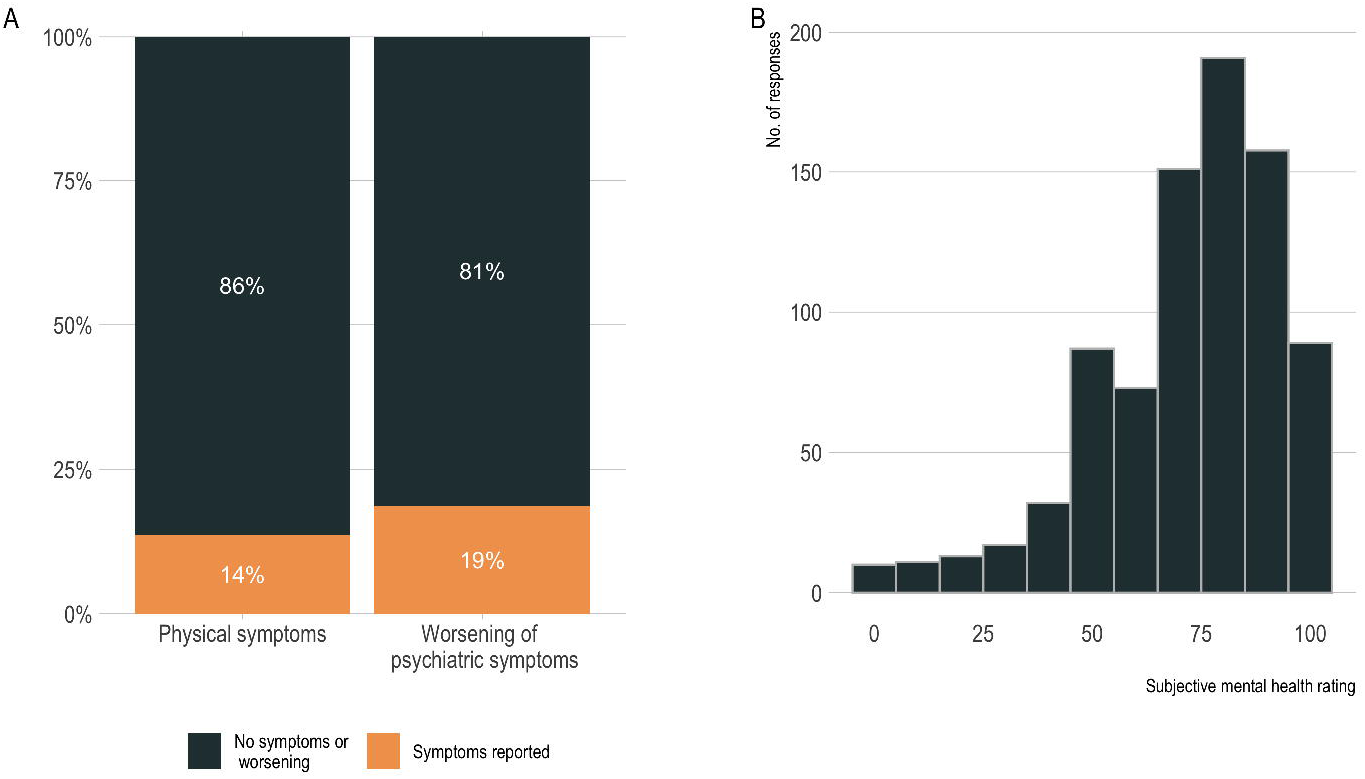
(A) Rates of respiratory symptoms, and rate of worsening of psychiatric symptoms as reported by patients. (B) Distribution of subjective mental health scores reported by patients.

